# BMI-related Genetic Factors and COPD Imaging Phenotypes

**DOI:** 10.1101/2024.08.16.24312135

**Authors:** Jingzhou Zhang, Matthew Moll, Catherine L. Debban, Brian D. Hobbs, Heena Rijhwani, George R. Washko, Bartolome R. Celli, Edwin K. Silverman, Per Bakke, Elizabeth C. Oelsner, R. Graham Barr, Alvar Agustí, Rosa Faner, Guy G. Bruselle, Stephen M. Humphries, David A. Lynch, Josée Dupuis, Ani W. Manichaikul, George T. O’Connor, Michael H. Cho

## Abstract

**Background:** While low body mass index (BMI) is associated with emphysema and obesity is associated with airway disease in chronic obstructive pulmonary disease (COPD), the underlying mechanisms are unclear.

**Methods:** We aggregated genetic variants from population-based genome-wide association studies to generate a polygenic score of BMI (PGS_BMI_). We calculated this score for participants from COPD-enriched and community-based cohorts and examined associations with automated quantification and visual interpretation of computed tomographic emphysema and airway wall thickness (AWT). We summarized the results using meta-analysis.

**Results:** In the random-effects meta-analyses combining results of all cohorts (n=16,349), a standard deviation increase of the PGS_BMI_ was associated with less emphysema as quantified by log-transformed percent of low attenuation areas ≤ 950 Hounsfield units (β= -0.062, *p*<0.0001) and 15^th^ percentile value of lung density histogram (β=2.27, *p*<0.0001), and increased AWT as quantified by the square root of wall area of a 10-mm lumen perimeter airway (β=0.016, *p*=0.0006) and mean segmental bronchial wall area percent (β=0.26, *p*=0.0013). For imaging characteristics assessed by visual interpretation, a higher PGS_BMI_ was associated with reduced emphysema in both COPD-enriched cohorts (OR for a higher severity grade=0.89, *p*=0.0080) and in the community-based Framingham Heart Study (OR for the presence of emphysema=0.82, *p*=0.0034), and a higher risk of airway wall thickening in the COPDGene study (OR=1.17, *p*=0.0023).

**Conclusions:** In individuals with and without COPD, a higher body mass index polygenic risk is associated with both quantitative and visual decreased emphysema and increased AWT, suggesting genetic determinants of BMI affect both emphysema and airway wall thickening.

## Introduction

Chronic obstructive pulmonary disease (COPD) is a heterogeneous condition, with emphysema and airway disease recognized as associated clinical phenotypes. In data-driven clustering studies of COPD phenotypes, emphysema- and airway-predominant disease axes and subtypes have been identified.^1,2^ On chest computed tomography (CT), emphysema is characterized by low attenuation areas and airway disease is associated with airway wall thickening. CT-based emphysema and airway wall thickness (AWT) measures are associated with airflow obstruction and mortality in people with and without COPD.^3–5^

Body mass index (BMI) varies greatly in people with COPD. Individuals with predominant emphysema often have lower BMI compared to those with predominant airway disease. This phenomenon was observed decades ago when the terms “pink puffer” and “blue bloater” were used to describe people with stereotypical emphysema and chronic bronchitis phenotypes, respectively.^6^ Using quantitative CT measures, lower BMI is associated with emphysema and higher fat mass is associated with higher AWT in individuals with COPD.^7,8^ However, the mechanisms underlying these observed associations are unclear.

Genetic variants are assigned at birth and thus less susceptible to later life confounding factors, offer an opportunity to identify causal disease mechanisms. Genetic factors strongly influence BMI, with an estimated heritability of 30-40% with common genetic variants explaining more than half of the heritability.^9^ Genome-wide association studies (GWAS) have identified hundreds of common genetic variants associated with BMI.^10^ The common variants most strongly associated with BMI, fat mass, and obesity are at the *FTO* (fat mass and obesity-associated) gene locus, both in general population samples and in patients with COPD.^11^ However, for complex traits such as BMI, the effect of an individual genetic variant tends to be small. A polygenic score (PGS) aggregating the effects of numerous variants across the entire genome can increase the power to detect genetic effects and clarify causal relationships.^12^

We hypothesized that genetic variants associated with higher BMI are associated with decreased emphysema and increased AWT. As emphysema and airway disease can precede the development of airflow obstruction and are associated with clinical outcomes in individuals with and without COPD,^3–5^ we tested our hypotheses among participants from COPD-enriched and community-based cohorts.

## Methods

### Study cohorts

We included participants who had available genotyping and chest CT data from the Genetic Epidemiology of COPD (COPDGene),^13^ the Evaluation of COPD Longitudinally to Identify Predictive Surrogate Endpoints (ECLIPSE),^14^ the National Emphysema Treatment Trial (NETT),^15^ the Genetics of Chronic Obstructive Lung Disease (GenKOLS),^16^ the Framingham Heart (FHS),^17,18^ and the Multi-Ethnic Study of Atherosclerosis (MESA) Lung studies,^19^ which have been previously described. The COPDGene, ECLIPSE, NETT, and GenKOLS enrolled participants with a smoking history and were enriched with COPD. In contrast, the FHS and MESA recruited community-dwelling participants without regard to a history of smoking or COPD. The COPDGene (non-Hispanic white, NHW, and African American, AA participants) and MESA (NHW, AA, Chinese, and Hispanic participants) were multi-ancestry studies, and the rest of the cohorts included predominantly NHW participants. Each study was approved by the Institutional Review Boards at participating institutions, and all participants provided written informed consent. Further details of study cohorts can be found in the Supplementary Methods.

### Polygenic score of BMI

To obtain a summative measure of genetic variants associated with BMI, we developed a polygenic score of BMI (PGS_BMI_) based on the GIANT Consortium and UK biobank BMI GWAS meta-analysis summary statistics (∼700,000 predominantly general population participants of European ancestry) using elastic net-based regression (lassosum v0.4.5).^10^ The details of the development of the PGS_BMI_ have been previously described.^20^ We calculated a PGS_BMI_ for each study participant as a weighted sum of variant effects using the lassosum- generated weights and the dosage of the BMI-increasing allele at each variant. We estimated the phenotypic variance explained by the PGS_BMI_ within each cohort using the R-squared method, and a randomly selected sample of unrelated participants was used for this estimation in FHS.

### Emphysema and airway wall thickness measures

We included quantitative and visual (qualitative) measures of emphysema and airway wall thickness (AWT) assessed on inspiratory CT. Based on CT radiodensity (in Hounsfield unit [HU]), emphysema was quantitatively evaluated using percent of low attenuation area ≤ -950 HU (%LAA-950), HU value at the 15th percentile of lung attenuation histogram (perc15), and lung volume-adjusted lung density (ALD).^21^ Based on visual assessment, the severity of overall, centrilobular, and paraseptal emphysema were qualitatively graded into ordinal categories.^22^ We estimated the longitudinal change in ALD using the average annual difference in ALD between the 5-year follow-up and baseline visits in COPDGene. AWT was quantitatively measured using the square root of wall area of a theoretical airway with a 10-mm lumen perimeter (Pi10, in mm) and the mean wall area percent of the segmental (WAP-seg) and subsegmental (WAP-subseg) bronchi,^23^ and qualitatively evaluated as the presence of airway wall thickening based on visual assessment.^24^ In addition to individual emphysema and AWT measures, we also included previously identified emphysema and airway disease axes and COPD imaging subtypes in COPDGene.^2,25^ Additional details about the imaging outcomes can be found in the Supplementary Methods.

### PGS_BMI_ association analyses

We standardized the PGS_BMI_ to a mean of 0 and a standard deviation (SD) of 1 within each cohort. As a result, the effect estimates for the PGS_BMI_ were at the scale of per one SD increase of the PGS_BMI_ in the association analyses. The %LAA-950 was natural log transformed to account for skewness. We determined five primary outcomes including log-transformed %LAA- 950 (log-%LAA-950), perc15, visual emphysema severity, Pi10, and WAP-seg which were available among most cohorts. We tested associations between the PGS_BMI_ and emphysema and AWT outcomes using multivariable linear, binary logistic, ordinal logistic (for visual emphysema severity), and multinomial logistic (for COPD imaging subtypes) regressions as appropriate for outcomes. All models were adjusted for age, sex, current smoking status, pack-years of smoking, CT scanner, and principal components of genetic ancestry. AWT outcomes were additionally adjusted for height to account for body size.^26^ To assess whether BMI-related variation in inspiration at the CT scanning substantially influenced the association between PGS_BMI_ and AWT outcomes, we conducted sensitivity analysis further adjusting for CT total lung capacity (available in COPDGene, GenKOLS, and FHS). Longitudinal change in ALD was further adjusted for baseline ALD and current smoking status at the follow-up visit. Family relatedness in FHS was accounted for using a kinship matrix. We performed inverse variance-weighted (IVW) random-effects meta-analyses to combine effect estimates among cohorts. For the association analyses between the PGS_BMI_ and the primary outcomes, we used a Bonferroni-corrected p-value (*p* < 0.05/5) to determine statistical significance. The rest of the analyses were considered exploratory and a p-value < 0.05 was used.

### Mendelian randomization analyses

We performed two-sample Mendelian randomization (MR) analyses with the “TwoSampleMR” R package to determine the effect of BMI (per SD increase) on quantitative imaging outcomes based on previous GWASs.^10,26^ We used variants strongly associated with BMI (*p*<5×10^-8^) and applied stringent LD clumping (r^2^<0.0001, 10000kb window). We used the IVW method as the primary analysis and performed sensitivity analyses using the weighted median and MR-Egger methods for significant IVW estimates. We assessed heterogeneity using Cochran’s Q statistic and assessed directional horizontal pleiotropy using the Egger intercept.

## Results

### Baseline characteristics

We included a total of 16,349 participants from COPD-enriched and community-based cohorts (Table 1). The community-based cohorts had an approximately even proportion of male and female participants, whereas the COPD-enriched cohorts had more males than females. There were substantial differences in smoking history and lung function among cohorts. The distribution of visual emphysema severity among cohorts (Table S1-S6), and visual airway wall thickening in COPDGene (Table S7) are shown in the Supplementary Results.

**Table 1.** Baseline characteristics of participants in each cohort (n=16,349).

| Variable | COPDGene<br>NHW<br>(n=5604) | COPDGene<br>AA<br>(n=1793) | ECLIPSE<br>(n=1596) | NETT<br>(n=374) | GenKOLS<br>(n=774) | FHS<br>(n=3167) | MESA<br>NHW<br>(n=1190) | MESA AA<br>(n=802) | MESA<br>Chinese<br>(n=406) | MESA<br>Hispanic<br>(n=643) |
| --- | --- | --- | --- | --- | --- | --- | --- | --- | --- | --- |
| Age (year) | 62.3 (13.5) | 53.2 (9.5) | 64.0 (10.0) | 68.0 (7.0) | 59.3 (15.8) | 49.0 (14.0) | 70.0 (15.0) | 69.0 (14.0) | 68.0 (15.0) | 67.0 (14.0) |
| Sex (male) | 2934 (52.4%) | 1032 (57.6%) | 1046 (65.5%) | 239 (63.9%) | 461 (59.6%) | 1652 (52.2%) | 589 (49.5%) | 366 (45.6%) | 207 (51.0%) | 307 (47.7%) |
| Current smoker | 2167 (38.7%) | 1420 (79.2%) | 550 (34.5%) | 0 (0%) | 356 (46.0%) | 254 (8.0%) | 83 (7.0%) | 91 (11.3%) | 14 (3.4%) | 40 (6.2%) |
| Ever smoker | 100% | 100% | 100% | 100% | 100% | 1520 (48.0%) | 721 (60.6%) | 478 (59.6%) | 117 (28.8%) | 330 (51.3%) |
| Pack-years of<br>smoking * | 42.0 (28.8) | 35.3 (24.3) | 45.0 (27.0) | 60.8 (40.0) | 22.3 (20.9) | 12.0 (20.3) | 16.5 (29.5) | 13.0 (26.6) | 8.5 (21.0) | 6.3 (16.7) |
| BMI (kg/m<sup>2</sup>) | 27.9 (7.5) | 27.8 (8.4) | 26.2 (6.8) | 25.0 (5.0) | 25.5 (5.3) | 27.0 (6.4) | 27.1 (6.5) | 29.5 (7.4) | 23.9 (4.2) | 28.8 (6.4) |
| FEV<sub>1</sub> % predicted | 77.9 (38.1) | 88.9 (32.6) | 46.3 (24.1) | 28.0 (10.0) | 76.1 (40.1) | 97.9 (17.4) | 93.5 (22.1) | 93.7 (24.8) | 94.3 (21.1) | 101.9 (23.5) |
| FEV<sub>1</sub>/FVC | 0.69 (0.24) | 0.76 (0.19) | 0.44 (0.17) | 0.32 (0.09) | 0.68 (0.26) | 0.76 (0.09) | 0.73 (0.10) | 0.76 (0.11) | 0.75 (0.09) | 0.77 (0.09) |
| %LAA-950 | 3.1 (8.6) | 1.2 (3.5) | 16.5 (18.1) | 14.8 (16.4) | 1.8 (8.4) | 2.1 (2.4) | 2.0 (3.4) | 1.3 (2.1) | 1.5 (2.6) | 0.8 (1.5) |
| perc15 | -924.0 (33.7) | -907.0 (36.9) | -953.0 (34.0) | -949.0 (24.0) | -912.0 (48.3) | -910.0 (27.8) | -918.5 (26.0) | -907.9 (31.6) | -916.6 (26.4) | -903.5 (33.8) |
| Pi10 | 3.66 (0.15) | 3.69 (0.16) | 4.39 (0.25) | 4.55 (0.58) | 4.82 (0.45) | 3.64 (0.25) | 4.27 (0.29) | 4.19 (0.27) | 4.18 (0.24) | 4.26 (0.27) |
| WAP-seg | 61.1 (4.0) | 61.4 (4.2) | 65.8 (5.5) | 73.2 (4.5) | 75.4 (3.9) | 52.9 (6.1) | 61.6 (4.4) | 61.1 (4.5) | 62.3 (4.2) | 61.9 (4.0) |

Continuous variables are presented with the median (interquartile range). Categorical variables are presented with count (percentage).
\* Among ever-smokers.
COPDGene = Genetic Epidemiology of COPD; ECLIPSE = Evaluation of COPD Longitudinally to Identify Predictive Surrogate Endpoints; NETT = National Emphysema Treatment Trial; GenKOLS = Genetics of Chronic Obstructive Lung Disease; FHS = Framingham Heart Study; MESA = Multi-Ethnic Study of Atherosclerosis; NHW = non-Hispanic white; AA = African American.
BMI = body mass index; FEV<sub>1</sub> = forced expiratory volume in 1s; FVC = forced vital capacity; %LAA-950 = percent of low attenuation area ≤ -950 Hounsfield units; perc15 = the 15<sup>th</sup> percentile of the lung attenuation histogram; Pi10 = the square root of wall area of a 10-mm lumen perimeter airway; WAP-seg = the mean wall area percent of segmental bronchi.

### Performance of the PGS_BMI_

The polygenic score of BMI (PGS_BMI_) was significantly associated with measured BMI in all cohorts (Table 2). However, the variance in BMI explained by the PGS_BMI_ differed substantially among cohorts, and in general, did not perform as well in the AA cohorts as in the NHW cohorts. Within MESA, the PGS_BMI_ had similar performance explaining BMI variation in the NHW, Chinese, and Hispanic participants.

**Table 2.**
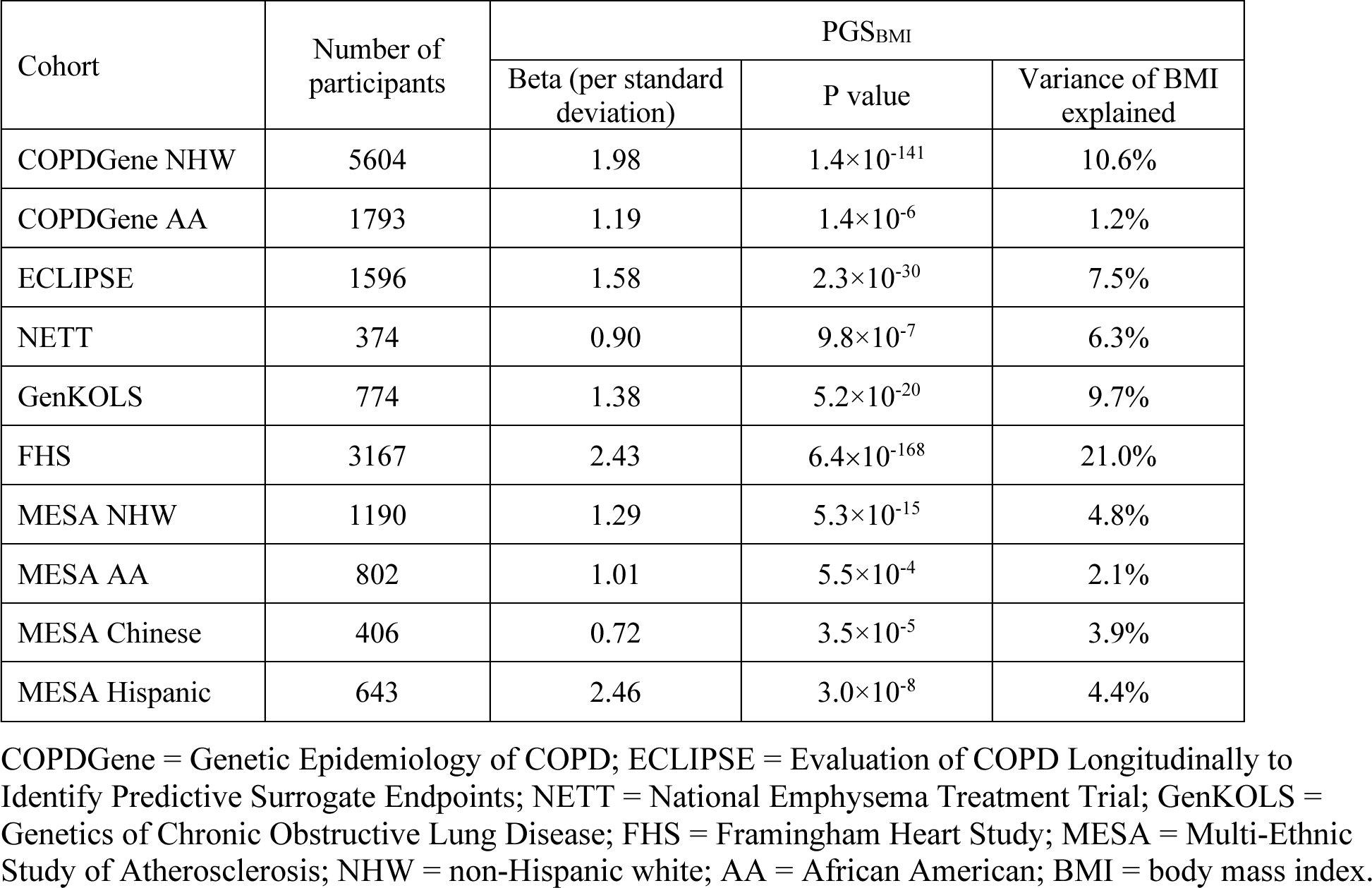
Associations of the polygenic score of BMI (PGS_BMI_) with measured BMI (kg/m^2^) in each study cohort.

### Associations between PGS_BMI_ and primary outcomes

In the random-effects meta-analysis of results across all cohorts (Figure 1), a standard deviation increase of the PGS_BMI_ was associated with decreased emphysema as indicated by a lower log-%LAA-950 (β= -0.062, *p*=7.6×10^-16^) and a higher perc15 (β=2.27, *p*=5.6×10^-8^), and increased AWT as indicated by a higher Pi10 (β=0.016, *p*=6.3×10^-4^) and WAP-seg (β=0.26, *p*=0.0013). In the meta-analysis of the COPD-enriched cohorts, a higher PGS_BMI_ was associated with a decreased risk of having a worse visual emphysema severity category (OR=0.89, *p*=0.0080). Consistent with this finding, in FHS participants, a higher PGS_BMI_ was associated with a lower risk for the presence (versus none) of visual emphysema (OR=0.82, *p*=0.0034). The results of the association between the PGS_BMI_ and AWT measures were similar after adjusting for CT lung volume in cohorts with this variable acquired (data not shown). We observed significant heterogeneity in the effect estimates among cohorts for the emphysema and AWT outcomes.

**Figure 1.**
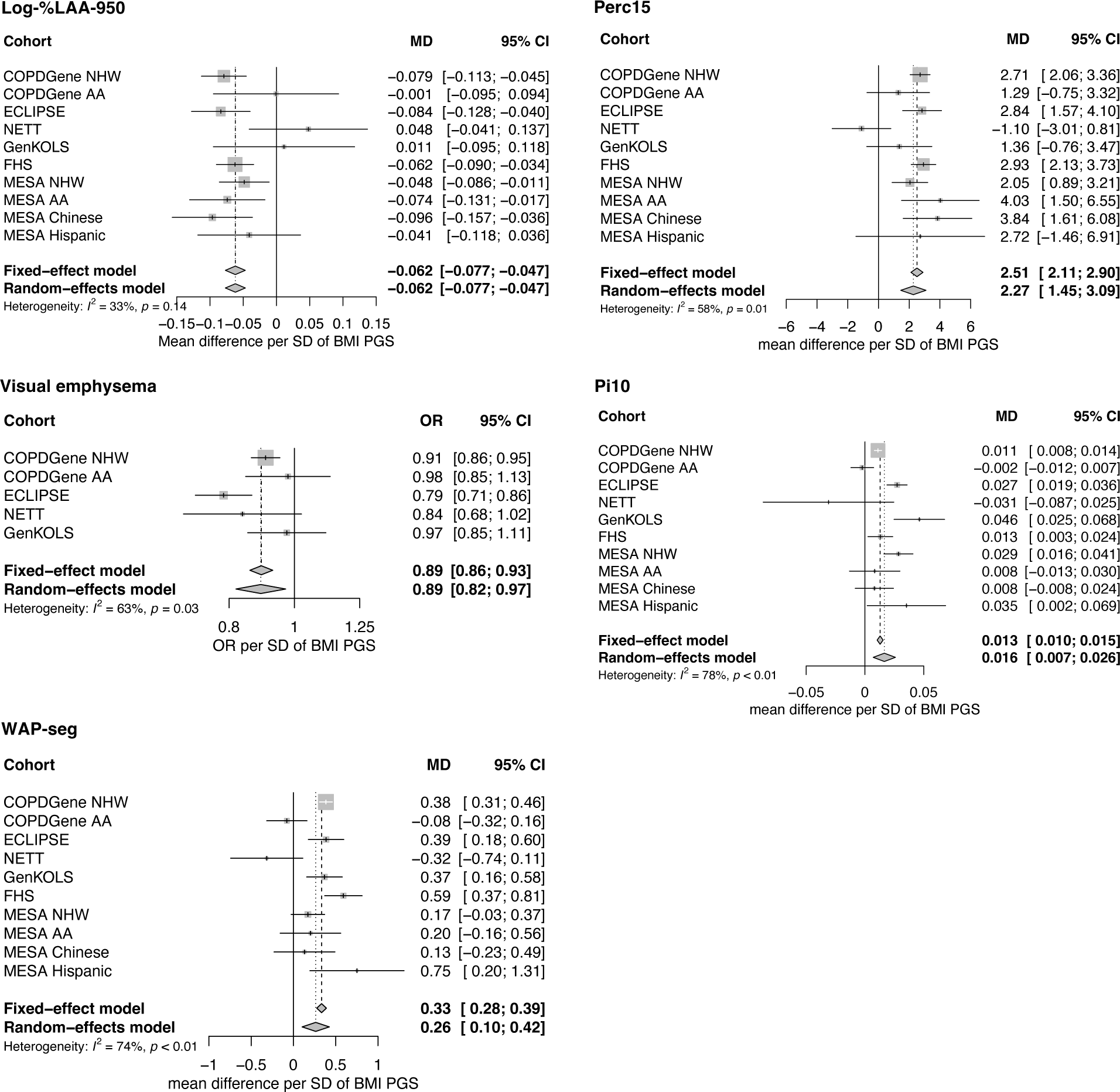
Associations of the body mass index (BMI) polygenic score (PGS) with computed tomographic emphysema and airway wall thickness measures among study cohorts. %LAA-950 = percent of low attenuation area ≤ -950 Hounsfield units; Perc15 = the 15^th^ percentile of the lung attenuation histogram; Pi10 = the square root of wall area of a 10-mm lumen perimeter airway; WAP-seg = the mean wall area percent of segmental bronchi; MD = mean difference; OR = odds ratio; SD = standard deviation.

### Associations between PGS_BMI_ and secondary outcomes

In the meta-analysis of results of COPDGene NHW and AA participants (Figure S1), a higher PGS_BMI_ was associated with a decreased risk of having a worse severity category of visual centrilobular (OR=0.91, *p*=1.2×10^-4^) and paraseptal (OR=0.91, *p*=3.4×10^-4^) emphysema, and an increased risk for the presence of visual airway wall thickening (OR=1.17, *p*=0.0023). In FHS participants, a higher PGS_BMI_ was associated with a lower risk for the presence of visual centrilobular (OR=0.81, *p*=0.048) and paraseptal (OR=0.83, *p*=0.032) emphysema. In the meta-analysis across the COPDGene and FHS (Figure S2), a higher PGS_BMI_ was associated with increased WAP of subsegmental bronchi (β=0.17, *p*=3.6×10^-4^).

In COPDGene, the PGS_BMI_ was associated with previously identified emphysema- and airway-predominant disease axes, and imaging subtypes (Table S8 and S9). A higher PGS_BMI_ was associated with a higher baseline lung volume-adjusted lung density; however, there was no significant association between the PGS_BMI_ and the longitudinal change in lung density (see details in Supplementary Results).

### Mendelian randomization analyses

The IVW MR analyses suggested a significant effect of genetically predicted BMI on perc15 (β per SD increase of BMI=4.66, *p*=8.5×10^-4^), Pi10 (β=0.026, *p*=6.8×10^-4^), and WAP-seg (β=0.80, *p*=3.1×10^-6^) (Table 3, MR scatterplots in Figures S3-S6). The effect of BMI on perc15 and WAP-seg remained significant using the weighted median and MR-Egger methods. We did not detect significant heterogeneity or directional horizontal pleiotropy of the instrumental variants’ effect.

**Table 3.** Causal effect estimates of BMI on emphysema and airway wall thickness measures.

| Outcome | MR methods | Variant number | Effect estimate* | p-value |
| --- | --- | --- | --- | --- |
| Log-%LAA-950 | IVW | 321 | -0.079 | 0.26 |
| perc15 | IVW | 321 | 4.66 | $8.5 \times 10^{-4}$ |
|  | Weighted median | 321 | 5.71 | 0.0090 |
|  | MR-Egger | 321 | 10.63 | 0.0025 |
| Pi10 | IVW | 321 | 0.026 | $6.8 \times 10^{-4}$ |
|  | Weighted median | 321 | 0.026 | 0.034 |
|  | MR-Egger | 321 | 0.019 | 0.31 |
| WAP-seg | IVW | 321 | 0.80 | $3.1 \times 10^{-6}$ |
|  | Weighted median | 321 | 0.89 | 0.0014 |
|  | MR-Egger | 321 | 1.20 | 0.0061 |

\* Effect estimate was based on per one standard deviation increase of BMI.
BMI = body mass index; MR = Mendelian randomization; IVW = inverse variance weighted; log-%LAA-950 = log transformed percent of low attenuation area ≤ -950 Hounsfield units; perc15 = the 15th percentile of the lung attenuation histogram; Pi10 = the square root of wall area of a 10-mm lumen perimeter airway; WAP-seg = the mean wall area percent of segmental bronchi.

## Discussion

In this multi-cohort analysis of adults from COPD-enriched and population-based cohorts, using computed tomographic (CT) measures, we found that a polygenic score of BMI (PGS_BMI_) was associated with quantitative and visual emphysema and airway measures. Consistent with the findings of measured BMI, the PGS_BMI_ was associated with decreased quantitative emphysema measures, less severe visually assessed emphysema, and decreased risk for emphysema-predominant disease axis and subtypes. Conversely, it was associated with increased quantitative airway wall thickness (AWT) measures and visual airway wall thickening, as well as a higher risk for airway-predominant disease axis and subtypes.

Automated quantification and visual interpretation of imaging characteristics capture distinct aspects and pathogenesis of emphysema and airway disease and provide complementary information on these phenotypes.^24,25^ Quantitative measures catch subtle radiodensity differences beyond visual detection and thus provide high measurement sensitivity. However, it is important to note that chest wall soft tissues, associated with BMI, could lead to variation in CT beam attenuation and thus affect the radiodensity of lung structures. In contrast, visual measures additionally capture morphological features, such as the size, type, and distribution of low attenuation areas, and can differentiate emphysematous lesions from other imaging abnormalities. Thus, visually assessed emphysema may be less susceptible to the BMI-related artifact compared to quantitative emphysema measures. Restrictive effects of BMI on the lungs may lead to variation in inspiration at CT scanning and affect airway lumen diameters. However, the association between the PGS_BMI_ and AWT measures remained when CT lung volume was additionally adjusted. Despite the potential biases introduced by BMI-related artifacts, these approaches helped to enhance the validity of our findings. Further studies using histological measures of emphysema and AWT may be warranted to confirm our findings; however, such studies are likely limited to a small sample size.

An association between low BMI and emphysema has been well described. However, cachexia is a well-known phenomenon in advanced COPD. Thus, whether low BMI itself is a risk factor for emphysema is not known. A previous study among participants with COPD have demonstrated associations of the *FTO* (fat mass and obesity-associated) gene-associated variants with BMI and emphysema;^11^ however whether these findings hold true for a set of BMI-related variants, for additional imaging phenotypes and in larger cohorts, is not known. Our finding of an inverse association between the PGS_BMI_ and emphysema suggests that a genetic predisposition to lower BMI is a risk factor for emphysema. BMI polygenic scores derived from GWASs of adult BMI explain BMI variation in early childhood,^27^ indicating that adult BMI-related genetic variants could exert effects on affected traits in a person’s early life. In humans, the number of alveoli increases substantially during the first two years of life and continues to increase at a slower rate until adolescence.^28^ Individuals with larger lungs have been shown to have lower lung density compared to those with smaller lungs, with implications of anthropometric traits affecting alveolar growth and structure.^29^ Recent studies showed that genes regulating organ size (e.g., skeletomuscular development) were enriched in lung development,^30^ and were shared between BMI and lung function.^31^ In extreme conditions, relatively young individuals who died of starvation or were severely malnourished demonstrated evidence of emphysema.^32,33^ Similarly, in mouse studies, calorie restriction resulted in alveolar loss and gas-exchange unit enlargement, consistent with histological emphysema, whereas subsequent refeeding led to alveolar regeneration.^34^ It was thus hypothesized that adaptive tissue degeneration of alveoli could occur in response to calorie restriction and decreased oxygen consumption. Biologic pathways that are activated in the setting of reduced calorie intake may be related to genetic variants leading to low BMI. An analysis of ECLIPSE showed that in people with COPD, emphysema was associated with low BMI and multi-organ extrapulmonary tissue loss, indicating a phenotype characterized by a defect in tissue repair and maintenance.^35^ The role of BMI-related genetics and related biological pathways in alveolar tissue growth, maintenance, and repair remains to be elucidated.

In contrast to emphysema, our finding of an association of a higher PGS_BMI_ with increased AWT implies a possible causal effect of obesity on bronchial wall thickening. Obesity is associated with systemic inflammation which affects multiple organs. In adults with asthma, obesity was associated with airway inflammation characterized by elevated bronchial submucosal eosinophils and sputum interleukin-5.^36^ In addition to the effect of circulating inflammatory cytokines released by remote adipocytes, adipose tissue within the airway wall may play a role in airway inflammation. Of note, a recent study of post-mortem lungs showed that adipose tissue was present in large airways, and was positively associated with BMI, number of inflammatory cells in the airway, and histological AWT.^37^ In animal studies, adipose cells were found in the airways of porcine lungs and functioned as a local source of inflammatory adipokines in the airway.^38^ Another study showed that obese adipocytes promoted inflammation of airway epithelial cells which was attenuated by weight loss in mice.^39^ Our findings raise the possibility that modification of BMI may affect airway disease among obese individuals, which needs to be further examined in clinical trials.

We used a PGS as the primary exposure given a higher phenotypic variance explained compared to an individual genetic variant. A PGS, comprising genetic variants fixed at conception, along with Mendelian randomization (MR) analyses, can provide support for causality.^12^ We studied variants identified from very large studies of BMI in population-based cohorts (not enriched for COPD or emphysema) and used MR methods that are robust to some forms of pleiotropy. However, we acknowledge that statements of causality in genetics can be complicated by effects that may occur outside the studied exposure and pleiotropy. It is worth noting that the effect size estimates from the PGS and the MR analyses are not directly comparable because the exposure variables are on different scales. Nonetheless, the primary goal of this study was not to quantify but to identify causal effects.

Additional limitations of the study include that different CT imaging protocols, data processing software, and visual emphysema scoring methods were utilized among study cohorts, which inevitably introduced heterogeneity and noise in the imaging measures. However, we still found significant results when combining estimates across cohorts using random-effects meta-analysis. BMI is a crude measure of adiposity and does not distinguish between the relative content or distribution of bone, muscle, or adipose tissue (i.e., body composition). Our PGS_BMI_ was derived from BMI GWASs using predominantly general population cohorts with European ancestry. This precluded our ability to examine the effect of variants of non-European ancestry, of rare allele frequency, or in a specific disease state. FHS contributed approximately 1% to the sample populations of the BMI GWAS where our PGS_BMI_ was derived, which may result in overfitting of the PGS_BMI_ in FHS participants. MESA had a considerable variant drop-out which may explain the relatively lower performance of the PGS_BMI_ among MESA cohorts. Our PGS_BMI_ did not perform as well in the African Americans as in the non-Hispanic white participants, which is consistent with the known issue of low portability of PGSs across ancestry.^40^ The PGS portability issue is thought primarily stemming from different linkage disequilibrium patterns and allele frequencies between populations, and highlights the importance of conducting multiancestry GWASs and constructing multi-ancestry PGSs in the future. Further, Chinese and Hispanic participants were only present in MESA with a relatively small sample size. The generalizability of our results to individuals of non-European ancestry needs to be confirmed.

To conclude, a higher BMI polygenic score is associated with decreased emphysema and increased airway wall thickness among persons with and without COPD. Our data suggest that genetic factors related to BMI also influence emphysema and airway disease, indicating potential shared genetic underpinnings of anthropometric and adiposity traits with COPD phenotypes.

Future studies integrating genetics of body composition traits beyond BMI, as well as multiomics data from blood and lung sources, are likely needed to further unravel the biological pathways underlying these associations. This exploration may lead to the discovery of novel clinical markers and potential druggable targets.

## Data Availability

All data produced in the present study are available upon reasonable request to the authors

