## Supplementary for "BMI-related Genetic Factors and COPD Imaging Phenotypes"

### Supplementary Methods

*Study cohorts*

COPDGene

The Genetic Epidemiology of COPD (COPDGene) study is a multicenter prospective observational study that enrolled smokers aged 45-80 years with a smoking history ≥10 pack-years at baseline (1). COPDGene aimed to enroll 10,000 former and current smoking participants from 21 clinical centers across the U.S., with a goal of 4,000 control participants ($FEV_1/FVC \geq 0.7$, $FEV_1 \geq 80\%$ predicted), 2,000 GOLD 1 and preserved ratio impaired spirometry (PRISm) participants, and 4,000 GOLD 2-4 participants. Non-Hispanic white (NHW) and African American (AA) participants were planned to be 2/3 and 1/3 of each group, respectively. Post-bronchodilator spirometry was available in COPDGene. Illumina (San Diego, CA) performed genotyping on the HumanOmniExpress array. Genotyping at the Z and S alleles was performed in all subjects. Subjects with severe alpha-1 antitrypsin deficiency were excluded. Imputation was performed by the Michigan Imputation Server to the Haplotype Resource Consortium and 1000 Genomes Phase I v3 Cosmopolitan reference panels, for NHW and AA participants, respectively. Variants with an r2 value of ≤ 0.3 were removed. We used baseline CT phenotypes analyzed on segmented lung images using VIDA Pulmonary Workstation, version 2.0 (VIDA Diagnostics, Coralville, Iowa). Automated airway segmentation and quantification were performed (2). For each bronchial tree, multiple parameters are calculated for third (segmental), fourth, fifth, and sixth generation bronchi, including wall area, lumen area, wall thickness, and luminal diameter. Visual emphysema analysis was performed by trained research analysts based on the Fleischner Society classification system (3), which has been previously described (4). Visual airway wall thickening was graded to none, possible, and definite.

ECLIPSE

The Evaluation of COPD Longitudinally to Identify Predictive Surrogate Endpoints (ECLIPSE) study was a case-control study of smokers with ≥ 10 pack years of smoking history, aged 40-75 years, and without other respiratory diseases (5). Post-bronchodilator spirometry was available in ECLIPSE. Genotyping was performed using the Illumina HumanHap 550 V3 (Illumina, San Diego, CA). Subjects and markers with a call rate of < 95% were excluded. Imputation was performed using the Michigan Imputation Server and Haplotype Resource Consortium2 reference panel. We used low-dose CT scans were performed at baseline for this analysis. All scans were performed using multidetector CT scans (GE Healthcare, Milwaukee, Wisconsin, or Siemens Healthcare, Erlangen, Germany) and images were reconstructed using 1.0 mm (Siemens) or 1.25mm (GE) contiguous slices and an intermediate spatial frequency reconstruction algorithm. All CT scans were analyzed at the University of British Columbia using Pulmonary Workstation 2.0 software (VIDA Diagnostics, Coralville, Iowa). Airways were segmented to the third (segmental) to fifth generation airways. Wall area percent was calculated using the mean value of measurements for selected segmental airways across all lobes. Visual analysis was performed by trained research analysts based on the Fleischner Society classification system (3).

NETT

The National Emphysema Treatment Trial (NETT) was a multicenter randomized clinical trial comparing lung-volume-reduction surgery and medical therapy for severe emphysema (6). All subjects in NETT were former smokers with severe COPD (FEV1 ≤45% predicted). Subjects with significant sputum production or bronchiectasis were excluded. Genotyping for NETT was performed using the Illumina Quad 610 array (Illumina, San Diego, CA). Imputation was performed using the Michigan Imputation Server and Haplotype Resource Consortium2 reference panel. NETT CT scans were performed on one of three types of scanners (General Electric, Fairfield, CT; Siemens, Malvern, PA; or Picker International, Toronto, ON, Canada) with a range of 2- to 8-mm slice thickness, with 75% of the scan data from 4 to 5 mm (7). Densitometric measures were performed with the Pulmonary Analysis Software Suite (PASS, Iowa City, Iowa). The extent of emphysema in six lung zones was visually inspected and qualitatively graded into scores of 1 to 4, representing emphysema area of no more than 25%, 26-50%, 51-75%, and more than 75% (8). Airway measurements were obtained using 3D Slicer (www.Slicer.org) and Airway Inspector (www.airwayinspector.org) at Brigham and Women's Hospital.

GenKOLS

The Genetics of Chronic Obstructive Lung Disease Study (GenKOLS) was a single-center case-control study based in Bergen, Norway, which included participants with a smoking history of > 2.5 pack years; severe alpha-1 antitrypsin deficiency and other lung diseases were excluded (9). The Regional Committee for Medical Research Ethics (REK Vest), the Norwegian Data Inspectorate, and the Norwegian Department of Health approved the case-control study. Written informed consent was obtained from all participants. Genotyping was performed using Illumina HumanHap 550 arrays (Illumina, San Diego, CA). Genotype imputation was the Michigan Imputation Server and Haplotype Resource Consortium reference panel. High-resolution CT chest scans were performed on a subset of the cohort using a GE LightSpeed Ultra. Images were analyzed at the James Hogg iCAPTURE Centre (Vancouver, BC, Canada). Emphysema was visually graded into qualitative scores from 0 to 5. Airways with an internal perimeter > 6mm were identified on the CT scans and measured using the Full Width at Half Maximum algorithm. Details of CT measures have been previously described (10).

FHS

The Framingham Heart Study (FHS) is a U.S. community-based cohort in Framingham, Massachusetts, established in 1948 (the Original cohort). The Offspring cohort began in 1971 and is comprised of children of the Original cohort and spouses of these children (11). The Third Generation (Gen3) cohort started in 2002 and is comprised of children from large Offspring cohort families (12). Pre-bronchodilator spirometry was performed at exams 5-9 of the Offspring cohort and exams 1-2 of the Gen3 cohort. The time intervals between exams were approximately 4-6 years. Genotyping was performed using the Affymetrix (Santa Clara, CA) GeneChip Human Mapping 500K Array Set, which was comprised of two arrays generating approximately 262,000 SNPs with Nsp arrays and 238,000 SNPs with Sty arrays. An additional Affymetrix 50K Array (HuGeneFocused50K) with gene-centric and coding SNPs was also genotyped for a total of approximately 550K SNPs. From 2002-2005, the Offspring and Gen3 cohorts underwent cardiac CT scans (Round 1 CT). Subsequently, from 2009-2011, chest CT scans (Round 2) which cover the entire lungs were performed. Non-contrast cardiac CT in supine using an 8-detector-row CT scanner (Lightspeed, GE Healthcare, Waukesha, WI) with 120 kV, 320−400 mA, a gantry rotation time of 0.5 s, and slice thickness of 2.5 mm was used for Round 1 CT (13). Non-contrast chest CT in supine at full inspiration using 64-detector-row CT scanner (Discovery, GE Healthcare, Waukesha, WI) with 120 kV and 300−400 mA (optimized with body weight), a gantry rotation time of 0.35 s and slice thickness of 0.63 mm was used for Round 2 CT (13). Emphysema was determined based on visual evaluation by three board-certified radiologists using a modified sequential reading method (14). The percent of low attenuation area ≤ -950 HU (%LAA-950) and HU value at the 15th percentile of lung attenuation histogram (perc15) on Round 1 CT were used in the current study. The lung density measures obtained through cardiac CT have been validated (15). Airway measures and visual interpretation of emphysema were available on Round 2 CT (n=2491) and were used for this study. Participants from the exams closest to Round 1 CT (Offspring Exam 7 and Gen3 Exam 1) and Round 2 CT (Offspring Exam 8 and Gen3 Exam 2) were used for corresponding imaging outcomes.

MESA

The Multi-Ethnic Study of Atherosclerosis (MESA) was a longitudinal study of subclinical cardiovascular disease and risk factors that predict progression to clinically overt cardiovascular disease or progression of the subclinical disease (16). MESA recruited 6,814 asymptomatic men and women aged 45-84 from six field centers across the United States in 2000-2002. In 2004-06, the MESA Lung Study recruited 3,965 participants sampled randomly from MESA participants. Minority race/ethnic groups were oversampled. The MESA Lung cohort comprises approximately 35% non-Hispanic whites, 24% African Americans, 23% Hispanics, and 18% Chinese Americans. Genotyping was performed at Affymetrix (Santa Clara, California, USA) and the Broad Institute of Harvard and MIT (Boston, Massachusetts, USA) using the Affymetrix Genome-Wide Human SNP Array 6.0, and these genome-wide array data were used to compute the BMI polygenic score in MESA. The MESA Lung Study performed full-lung CT scans on six MDCT scanners for over 3,200 participants in 2010-2012 following the MESA Lung/SPIROMICS CT protocol (17). The MESA Lung Study 2010-2012 was used for this study.

*Statistical analysis*

Visual emphysema severity category in COPDGene (Table S1) and ECLIPSE (Table S2) was used as the ordinal outcomes for ordinal logistic regression. In the NETT, we categorized the average visual emphysema scores using cutoffs of 2 and 3 to further categorize visual emphysema severity into mild to moderate, severe, and very severe (Table S3), which was used as the ordinal outcome. For GenKOLS, we used cutoffs of 1, 2, 3, 4, and 5 to categorize the visual emphysema scores into the ordinal severity category (Table S4). In FHS, given most participants had none or mild visually assessed emphysema, we examined visual emphysema as a binary outcome (presence versus none) using logistic regression analysis. For the presence of visual airway wall thickening in COPDGene (Table S7), we examined a binary outcome (definite versus none) using logistic regression analysis.

### Supplementary Results

In the COPDGene NHW participants, a standard deviation increase of the $PGS_{BMI}$ was associated with previously identified emphysema-predominant (β= -0.078, $p$=2.7×10$^{-11}$) and airway-predominant (β=0.11, $p$=7.1×10$^{-16}$) disease axes. We did not observe a significant association of the $PGS_{BMI}$ with the emphysema-predominant (β= -0.028, $p$=0.39) or airway-predominant (β= -0.029, $p$=0.44) disease axis in the COPDGene AA participants.

A higher $PGS_{BMI}$ was associated with increased lung volume-adjusted lung density (in Hounsfield unit) in the COPDGene NHW (β=3.05, $p$=4.6×10$^{-25}$) and AA (β=2.39, $p$=0.0068) participants. We did not observe a significant association of the $PGS_{BMI}$ with the average annual change in lung density in the COPDGene NHW (β=0.038, $p$=0.35) or AA (β=0.052, $p$=0.72) participants.

### Supplementary Tables

Table S1. Visual emphysema severity classification in the Genetic Epidemiology of COPD (COPDGene) non-Hispanic white and African American participants.

| Emphysema severity category | Number of participants | |
|---|---|---|
| | Non-Hispanic white | African American |
| Normal | 1844 | 612 |
| Trace centrilobular emphysema | 897 | 374 |
| Mild centrilobular emphysema | 1026 | 398 |
| Moderate centrilobular emphysema | 904 | 252 |
| Confluent emphysema | 681 | 127 |
| Advanced destructive emphysema | 252 | 30 |

Table S2. Visual emphysema severity scores in the Evaluation of COPD Longitudinally to Identify Predictive Surrogate Endpoints (ECLIPSE) participants.

| Emphysema severity category | Number of participants |
|---|---|
| Not affected | 89 |
| Trivial | 404 |
| Mild | 339 |
| Moderate | 294 |
| Severe | 289 |
| Very severe | 128 |

Table S3. Visual emphysema severity category in the National Emphysema Treatment Trial (NETT) participants.

| Emphysema severity category | Number of participants |
|---|---|
| Mild to moderate | 46 |
| Severe | 155 |
| Very severe | 173 |

Table S4. Visual emphysema severity category in the Genetics of Chronic Obstructive Lung Disease Study (GenKOLS) participants.

| Emphysema severity category | Number of participants |
|---|---|
| Not affected | 235 |
| Trivial | 274 |
| Mild | 120 |
| Moderate | 70 |
| Severe | 50 |
| Very severe | 18 |

Table S5. Visual emphysema classification in the Framingham Heart Study participants*.

| Emphysema classification | Number of participants |
|---|---|
| None | 2161 |
| CLE/CLE-dominant | 161 |
| Mixed CLE and PSE | 18 |
| PSE/PSE-dominant | 106 |
| Apical | 45 |

* Based on Round 2 CT.
CLE = centrilobular emphysema; PSE=paraseptal emphysema.

Table S6. Visual paraseptal emphysema severity classification in the Genetic Epidemiology of COPD (COPDGene) non-Hispanic white and African American participants.

| Emphysema severity category | Number of participants | |
|---|---|---|
| | Non-Hispanic white | African American |
| None | 3103 | 788 |
| Mild | 1390 | 557 |
| Substantial | 1111 | 448 |

Table S7. Visual airway wall thickening in the Genetic Epidemiology of COPD (COPDGene) non-Hispanic white and African American participants.

| Emphysema severity category | Number of participants | |
|---|---|---|
| | Non-Hispanic white | African American |
| None | 660 | 191 |
| Possible | 2609 | 977 |
| Definite | 2335 | 625 |

Table S8. Associations between the $PGS_{BMI}$ and COPD imaging subtypes in the Genetic Epidemiology of COPD (COPDGene) non-Hispanic white participants.

| Imaging subtype | Number of participants | OR (95% CI) * | P value |
|---|---|---|---|
| No CT imaging abnormality (reference) | 1470 | 1.00 | - |
| Paraseptal emphysema | 1106 | 0.84 (0.77, 0.91) | $6.2 \times 10^{-5}$ |
| Bronchial disease | 435 | 1.12 (1.00, 1.25) | 0.048 |
| Small airway disease | 253 | 0.90 (0.79, 1.03) | 0.14 |
| Mild emphysema | 1122 | 0.95 (0.88, 1.04) | 0.26 |
| Upper lobe predominant centrilobular emphysema | 218 | 0.99 (0.85, 1.15) | 0.90 |
| Lower lobe predominant centrilobular emphysema | 43 | 0.83 (0.62, 1.12) | 0.22 |
| Diffuse centrilobular emphysema | 538 | 0.87 (0.78, 0.97) | 0.011 |
| Visual without quantitative emphysema | 303 | 0.96 (0.84, 1.09) | 0.51 |
| Quantitative without visual emphysema | 99 | 0.77 (0.62, 0.96) | 0.021 |

$PGS_{BMI}$ = polygenic score of body mass index.
* Per standard deviation increase of the $PGS_{BMI}$

Table S9. Associations between the $PGS_{BMI}$ and COPD imaging subtypes in the Genetic Epidemiology of COPD (COPDGene) African American participants.

| Imaging subtype | Number of participants | OR (95% CI) * | P value |
|---|---|---|---|
| No CT imaging abnormality (reference) | 605 | 1.00 | - |
| Paraseptal emphysema | 448 | 0.92 (0.76, 1.10) | 0.35 |
| Bronchial disease | 146 | 0.87 (0.72, 1.04) | 0.12 |
| Small airway disease | 74 | 0.80 (0.62, 1.03) | 0.08 |
| Mild emphysema | 284 | 0.92 (0.78, 1.09) | 0.34 |
| Upper lobe predominant centrilobular emphysema | 47 | 0.98 (0.72, 1.33) | 0.88 |
| Lower lobe predominant centrilobular emphysema | 5 | 0.80 (0.37, 1.72) | 0.56 |
| Diffuse centrilobular emphysema | 72 | 0.86 (0.65, 1.14) | 0.29 |
| Visual without quantitative emphysema | 103 | 1.00 (0.80, 1.27) | 0.97 |
| Quantitative without visual emphysema | 6 | 0.83 (0.34, 2.00) | 0.68 |

$PGS_{BMI}$ = polygenic score of body mass index; CLE=centrilobular emphysema.
* Per standard deviation increase of the $PGS_{BMI}$

### Supplementary Figures

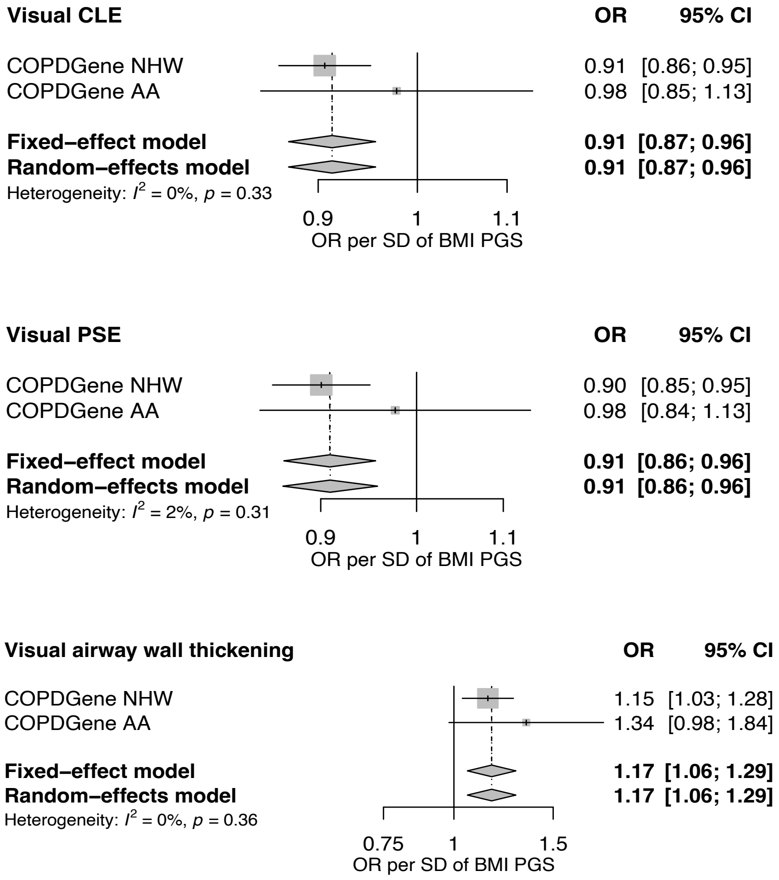

Figure S1. Associations of the body mass index (BMI) polygenic score (PGS) with visual centrilobular emphysema (CLE), paraseptal emphysema (PSE), and the presence of airway wall thickening among the Genetic Epidemiology of COPD (COPDGene) non-Hispanic white (NHW) and African American (AA) participants.

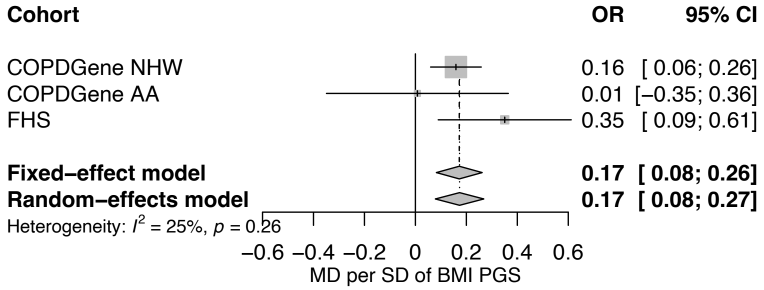

Figure S2. Associations of the body mass index (BMI) polygenic score (PGS) with the mean wall area percent of subsegmental bronchi among the Genetic Epidemiology of COPD (COPDGene) non-Hispanic white (NHW) and African American (AA) and Framingham Heart Study (FHS) participants.

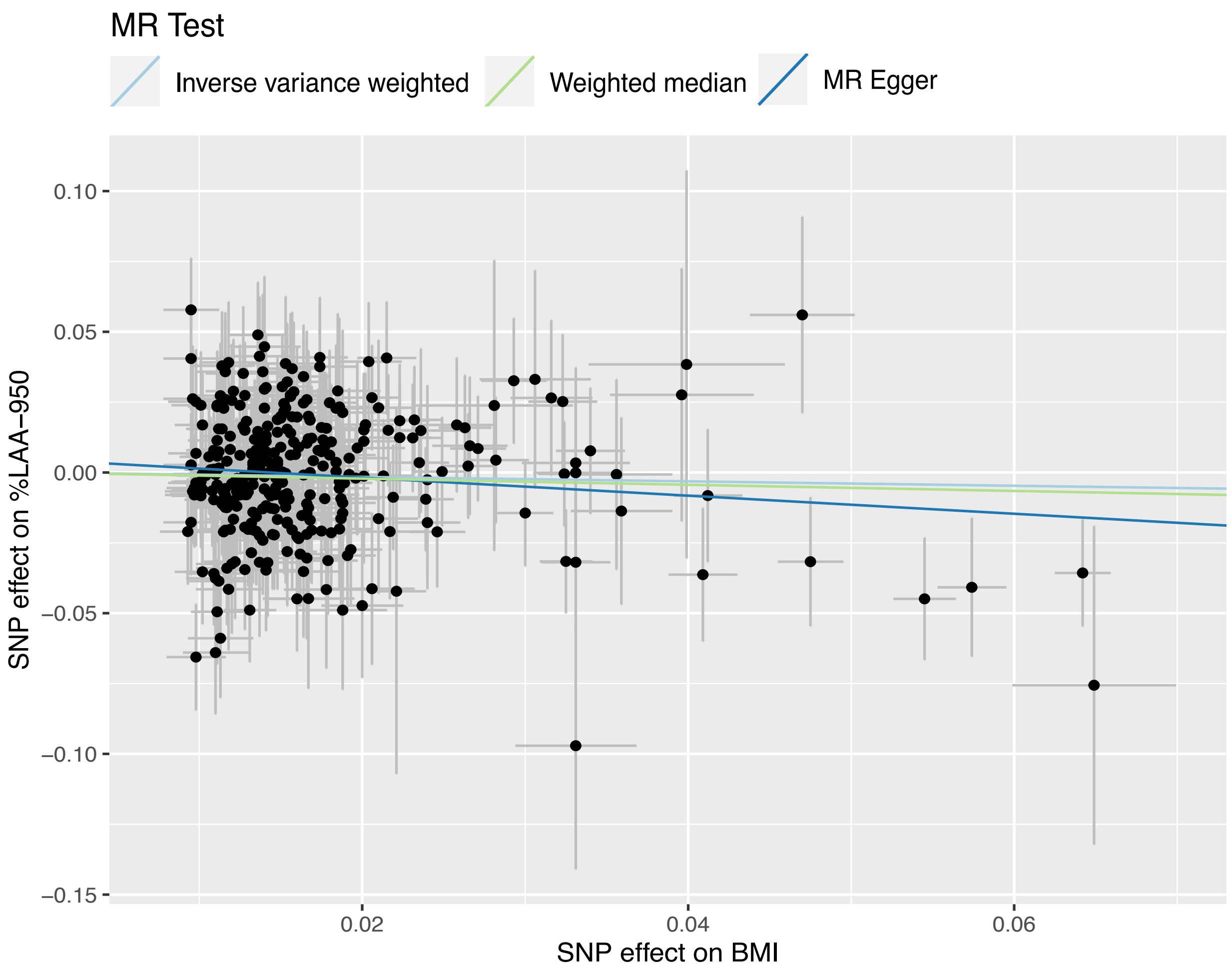

Figure S3. Scatterplots of Mendelian randomization instrumental single nucleotide polymorphism (SNP) effects on body mass index (BMI) and log-transformed percent of low attenuation area ≤ -950 Hounsfield units (%LAA-950). To facilitate visual interpretation, the SNP-BMI association estimates were uniformly made positive by convention, with corresponding changes for the BMI-outcome association estimates being flipped as appropriate.

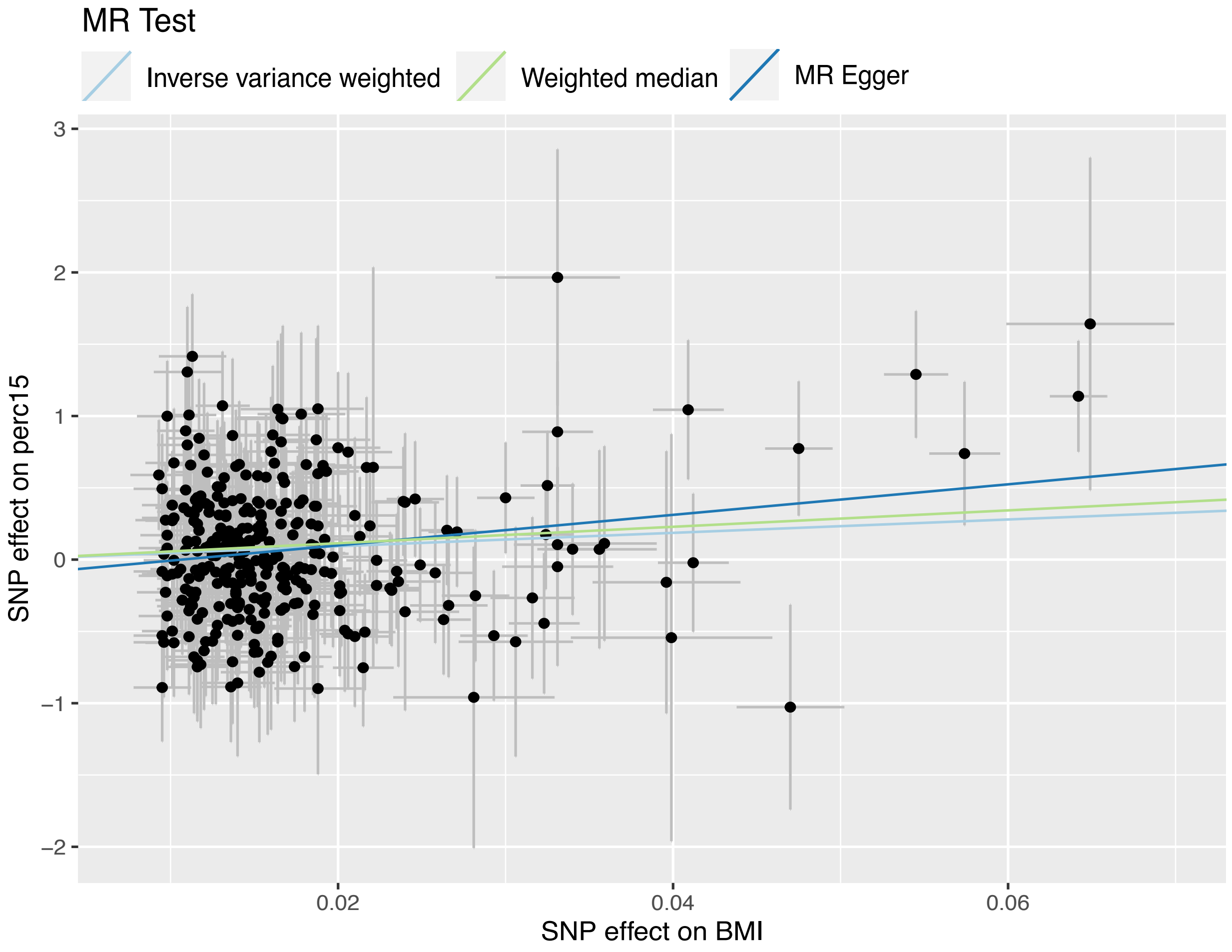

Figure S4. Scatterplots of Mendelian randomization instrumental single nucleotide polymorphism (SNP) effects on body mass index (BMI) and the 15$^{th}$ percentile of the lung attenuation histogram (perc15). To facilitate visual interpretation, the SNP-BMI association estimates were uniformly made positive by convention, with corresponding changes for the BMI-outcome association estimates being flipped as appropriate.

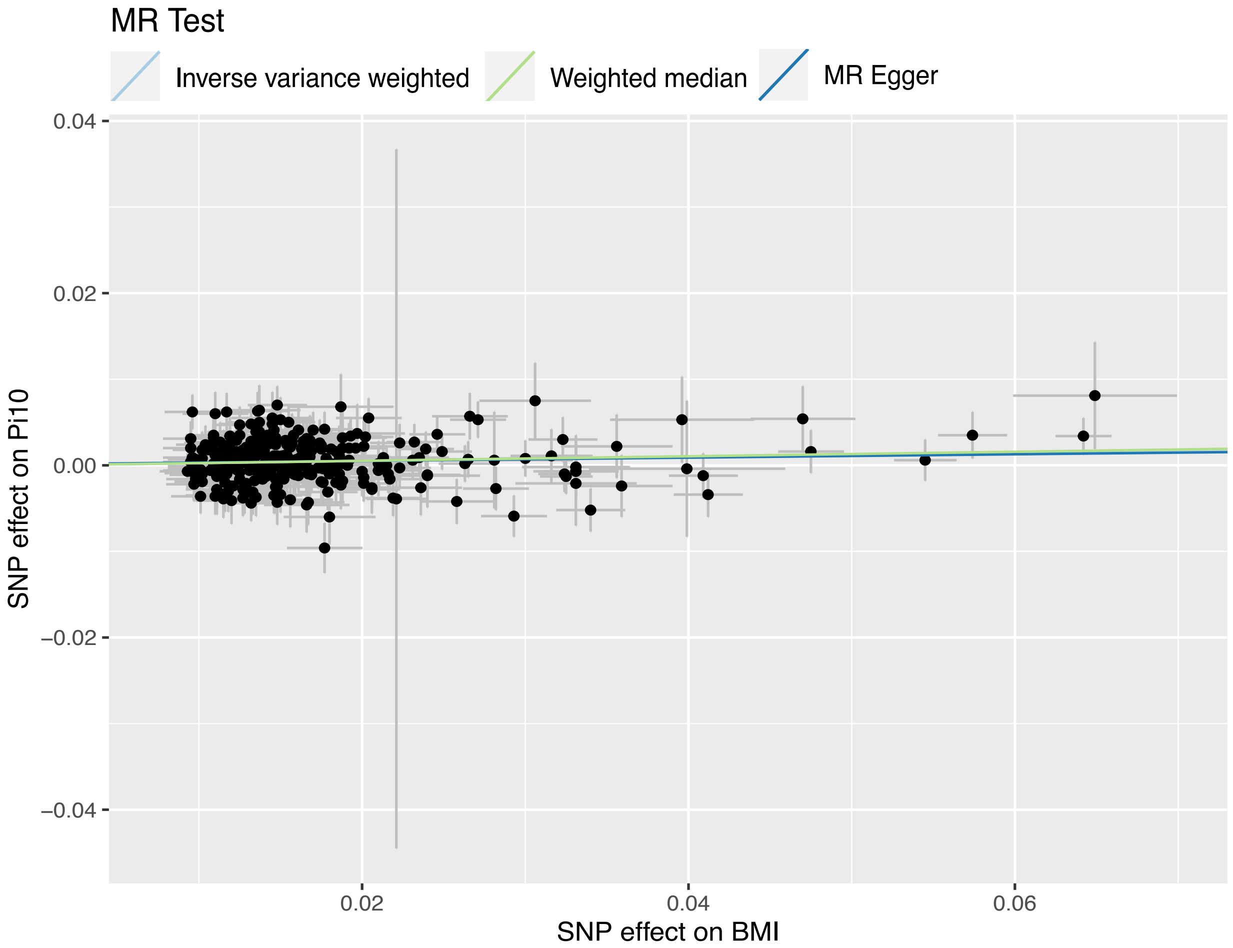

Figure S5. Scatterplots of Mendelian randomization instrumental single nucleotide polymorphism (SNP) effects on body mass index (BMI) and the square root of wall area of a 10-mm lumen perimeter (Pi10). To facilitate visual interpretation, the SNP-BMI association estimates were uniformly made positive by convention, with corresponding changes for the BMI-outcome association estimates being flipped as appropriate.

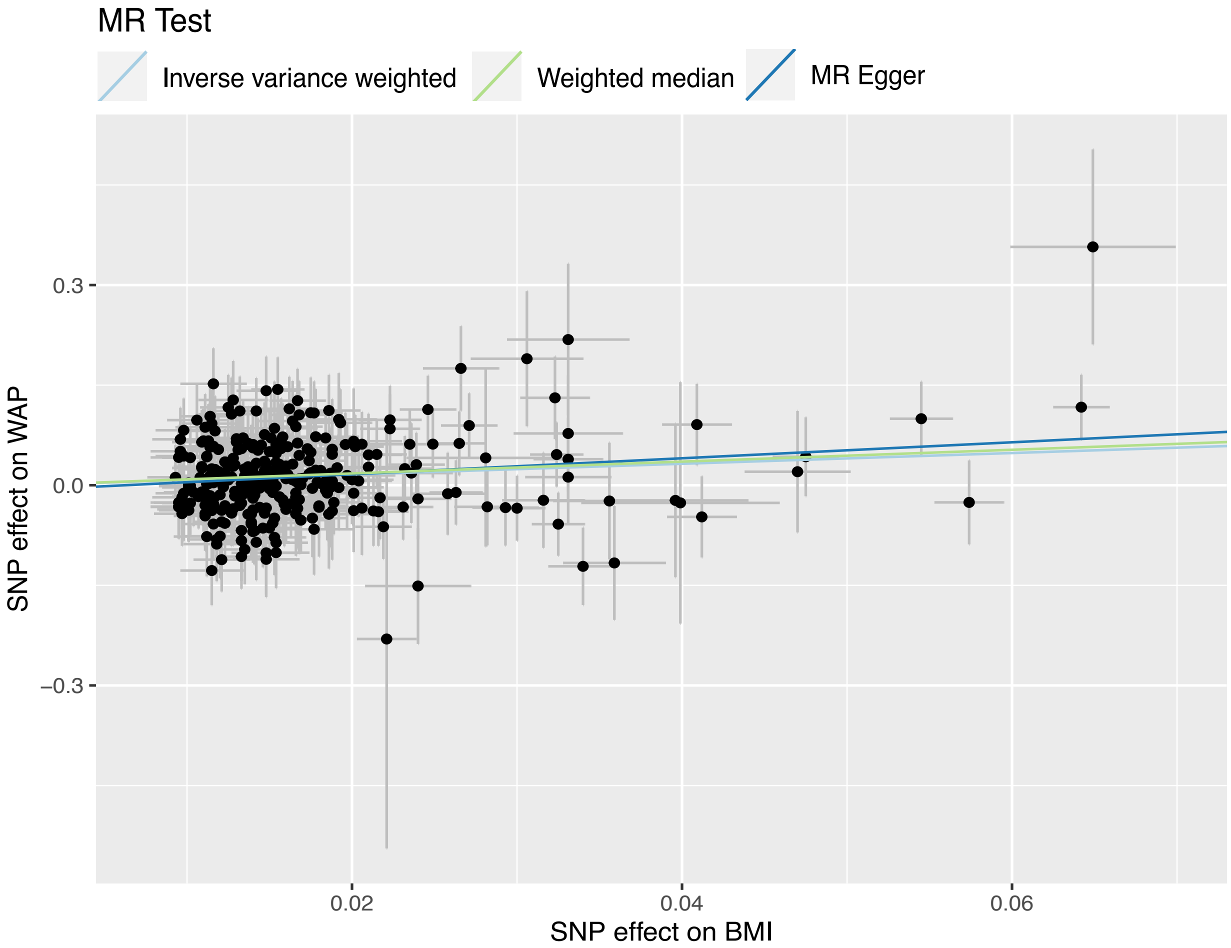

Figure S6. Scatterplots of Mendelian randomization instrumental single nucleotide polymorphism (SNP) effects on body mass index (BMI) and the mean wall area percent (WAP) of segmental bronchi. To facilitate visual interpretation, the SNP-BMI association estimates were uniformly made positive by convention, with corresponding changes for the BMI-outcome association estimates being flipped as appropriate.

**Funding and acknowledgments**

COPDGene was supported by NHLBI grants U01 HL089897 and U01 HL089856 and by NIH contract 75N92023D00011. The COPDGene study (NCT00608764) has also been supported by the COPD Foundation through contributions made to an Industry Advisory Committee that has included AstraZeneca, Bayer Pharmaceuticals, Boehringer-Ingelheim, Genentech, GlaxoSmithKline, Novartis, Pfizer, and Sunovion.

The ECLIPSE study (NCT00292552; GSK code SCO104960) was funded by GlaxoSmithKline.

The Framingham Heart Study is conducted and supported by the National Heart, Lung, and Blood Institute (NHLBI) in collaboration with Boston University (Contract No. N01-HC-25195, HHSN268201500001I and 75N92019D00031).

The Multi-Ethnic Study of Atherosclerosis (MESA) Lung Study was supported by NHLBI (NIH) grants R01-HL077612, R01-HL093081, and RC1-HL100543. Whole genome sequencing (WGS) for the Trans-Omics in Precision Medicine (TOPMed) program was supported by NHLBI. MESA and the MESA SHARe project are conducted and supported by NHLBI in collaboration with MESA investigators. The provision of genotyping data was supported in part by the National Center for Advancing Translational Sciences, CTSI grant UL1TR001881, and the National Institute of Diabetes and Digestive and Kidney Disease Diabetes Research Center (DRC) grant DK063491 to the Southern California Diabetes Endocrinology Research Center. Funding for SHARe genotyping was provided by NHLBI Contract N02-HL-64278. WGS for "NHLBI TOPMed: Multi-Ethnic Study of Atherosclerosis (MESA)" (phs001416.v3.p1) was performed at the Broad Institute of MIT and Harvard (3U54HG003067-13S1). Centralized read mapping and genotype calling, along with variant quality metrics and filtering were provided by the TOPMed Informatics Research Center (3R01HL-117626-02S1). Phenotype harmonization, data management, sample-identity QC, and general study coordination, were provided by the TOPMed Data Coordinating Center (3R01HL-120393-02S1). Support for the Multi-Ethnic Study of Atherosclerosis (MESA) projects are conducted and supported by the National Heart, Lung, and Blood Institute (NHLBI) in collaboration with MESA investigators. Support for MESA is provided by contracts 75N92020D00001, HHSN268201500003I, N01-HC-95159, 75N92020D00005, N01-HC-95160, 75N92020D00002, N01-HC-95161, 75N92020D00003, N01-HC-95162, 75N92020D00006, N01-HC-95163, 75N92020D00004, N01-HC-95164, 75N92020D00007, N01-HC-95165, N01-HC-95166, N01-HC-95167, N01-HC-95168, N01-HC-95169, UL1-TR-000040, UL1-TR-001079, UL1-TR-001420, UL1TR001881, DK063491, and R01HL105756. The authors thank the other investigators, the staff, and the participants of the MESA study for their valuable contributions. A full list of participating MESA investigators and institutes can be found at http://www.mesa-nhlbi.org.
